# Impact of Age-related change in Caval Flow Ratio on Hepatic Flow Distribution in Fontan

**DOI:** 10.1101/2023.09.06.23295166

**Authors:** V. Govindarajan, L. Marshall, A. Sahni, M. Cetatoiu, E. Eickhoff, J. Davee, N. St. Clair, N. Schulz, D.M. Hoganson, P. E. Hammer, S. Ghelani, A. Prakash, P. J. del Nido, R.H. Rathod

**Author notes:** Address of Correspondence Vijay Govindarajan, PhD, Department of Cardiovascular Surgery Boston Children’s Hospital, 300 Longwood Avenue Boston, MA 02115.

## Abstract

**Background:** The Fontan operation is a palliative technique for patients born with single ventricle heart disease. The superior vena cava (SVC), inferior vena cava (IVC), and hepatic veins are connected to the pulmonary arteries in a total cavopulmonary connection by an extracardiac (EC) conduit or a lateral tunnel (LT) connection. A balanced hepatic flow distribution (HFD) to both lungs is essential to prevent pulmonary arteriovenous malformations and cyanosis. HFD is highly dependent on the local hemodynamics.

**Objective:** The effect of age-related changes in caval inflows on HFD was evaluated using cardiac MRI (CMR) data and patient-specific computational fluid dynamics (CFD) modeling.

**Methods:** SVC and IVC flow from 414 Fontan patients were collected to establish a relationship between SVC:IVC flow ratio and age. CFD modeling was performed in 60 (30 EC and 30 LT) patient models to quantify the HFD that corresponded to patient ages of 3, 8, and 15 years, respectively.

**Results:** SVC:IVC flow ratio inverted at ∼8 years of age, indicating a clear shift to lower body flow predominance. Our data showed that variation of HFD in response to age-related changes in caval inflows (SVC:IVC = 2,1, and 0.5 corresponded to ages 3, 8, and 15+ respectively) was not significant for EC but statistically significant for LT cohorts. For all three caval inflow ratios, a positive correlation existed between the IVC flow distribution to both the lungs and the HFD. However, as the SVC:IVC ratio changed from 2→0.5 (age 3→15+), the correlation’s strength decreased from 0.87→0.64, due to potential flow perturbation as IVC flow momentum increased.

**Conclusion:** Our analysis provided quantitative insights into the impact of the changing caval inflows on Fontan’s long-term HFD, highlighting the importance of including SVC:IVC variations over time to understand Fontan’s long-term hemodynamics. These findings broaden our understanding of Fontan hemodynamics and patient outcomes.

**Clinical Perspective:** With improvement in standard of care and management of single ventricle patients with Fontan physiology, the population of adults with Fontan circulation is increasing. Consequently, there is a clinical need to comprehend the impact of patient growth on Fontan hemodynamics. Using CMR data, we were able to quantify the relationship between changing caval inflows and somatic growth. We then used patient-specific computational flow modeling to quantify how this relationship affected the distribution of long-term hepatic flow in extracardiac and lateral tunnel Fontan types. Our findings demonstrated the significance of including SVC:IVC changes over time in CFD modeling to learn more about the long-term hemodynamics of Fontan. Fontan surgical approaches are increasingly planned and optimized using computational flow modeling. For a patient undergoing a Fontan procedure, the workflow presented in this study that takes into account the variations in Caval inflows over time can aid in predicting the long-term hemodynamics in a planned Fontan pathway.

## Introduction

A successful Fontan procedure offers complete redirection of systemic venous return, improved arterial saturations, and decreased chronic ventricular volume overload, thereby improving longevity for many patients with single ventricle heart disease (SVHD)^1^. The Fontan circulation is the result of a total cavopulmonary connection between the superior vena cava (SVC), inferior vena cava (IVC), and the hepatic veins to the pulmonary arteries (PA) typically achieved either through an extracardiac (EC) conduit or a lateral tunnel (LT) connection. Although the Fontan procedure is lifesaving for patients with SVHD, this “unnatural” physiology can lead to serious early and late complications in many patients. These comorbidities include obligatory central venous hypertension, pulmonary arteriovenous malformations (PAVM) leading to cyanosis, exercise intolerance, ventricular dysfunction, arrhythmia, thrombus formation, stroke, protein losing enteropathy, exercise intolerance, pleural effusions, and hepatic and/or renal dysfunction ^1–3^.

Pulmonary arteriovenous malformations (PAVM) are an important cause of progressive cyanosis in patients with a Fontan circulation. PAVMs are abnormal communications that develop between the pulmonary artery and pulmonary veins that bypasses the normal capillary bed impacting gas exchange^4^ leading to progressive cyanosis and exercise intolerance^4, 5^. Complete resolution of PAVM have been achieved post-liver transplantation in the setting of severe liver disease, post-Fontan completion, and after redirection of hepatic venous blood to the affected lung^6–9^. These studies stress the importance of symmetric hepatic blood flow to both lungs for the prevention of PAVMs. Symmetric delivery of hepatic venous blood to each lung is dependent on local blood flow dynamics, Fontan geometries, and caval blood flow interactions^10–13^.

Cardiac magnetic resonance (CMR) imaging allows for the assessment of vascular anatomy and flow and, when combined with computational fluid dynamics (CFD), can offer patient-specific biomechanical insights improving our quantitative understanding of the complex hemodynamics in Fontan. Numerous CMR-CFD-based studies have been conducted to gain quantitative insights to evaluate Fontan blood flow dynamics^14–20^. These studies range from estimating the energy loss across the Fontan pathway^15, 18, 21^, determining optimal hemodynamics^20^, characterizing hepatic venous flow^12^, and predicting the non-Newtonian effects of blood in Fontan flow^22^. CFD-based modeling frameworks are now being extended to perform virtual surgeries to personalize and improve surgical predictability^10, 23–25^. While these studies have offered important insights into Fontan hemodynamics, shortcomings included small sample sizes, exclusion of hepatic inflow resulting in a limited understanding of local flow dynamics in hepatic-IVC-Fontan confluence, and simulation of only single flow states.

The goal of this study is to perform a comprehensive analysis of hepatic flow distribution (HFD) in a large cohort of LT and EC Fontan patients and quantitatively evaluate the impact of growth-related changes in caval inflow on hemodynamics with particular emphasis on HFD. To this end, we aim to 1) describe the quantitative relationship between the change in SVC-to-IVC inflow (interchangeably referred to as caval inflow) ratio over age, 2) perform CMR-based patient CFD modeling at different SVC-to-IVC flow ratios to quantitatively evaluate the variability of HFD over time, and 3) evaluate the correlations between IVC flow distribution to branch PAs and HFD supported by momentum analysis to delineate the impact of increasing IVC flow on HFD.

## Material and Methods

### Patient selection – Inclusion and exclusion criteria

Patients after the Fontan operation who had a CMR at Boston Children’s Hospital from 2003 to 2023 were retrospectively reviewed. Inclusion criteria included patients who had an EC or LT Fontan procedure as their primary surgery with non-Fenestrated Fontan or status-post closure. Patients were excluded if they had an interrupted IVC, bilateral bidirectional Glenn procedure, prior history of PAVM, history of thrombosis, or aortopulmonary collateral flow > 40%. This study was approved by the institutional review board at Boston Children’s Hospital.

A random subset of 30 patients with an EC Fontan and 30 patients with LT Fontan were chosen for computational flow modeling. To be included for CFD modeling, patients had to have minimal imaging artifacts, 3D isotropic SSFP imaging, and 2D flow assessment in the SVC, IVC, and branch PAs. Patient specific demographic information and CMR derived flow data are provided in supplementary tables 1 and 2.

### Patient-specific anatomy reconstruction from CMR images

Figure 1 illustrates the overall patient-specific workflow for evaluating Fontan flow dynamics for this study. The SVC, IVC, branch PAs, Fontan pathway, and major hepatic vessels were segmented using a threshold appropriate for the blood pool contrast from the 3D isotropic imaging sequences. Segmentation was performed using Mimics 25.0 (Materialise NV, Leuven, Belgium). The geometries were smoothed, and the mesh was constructed using 3-Matic Medical (Materialise NV, Leuven, Belgium) (Figure 1). The STL mesh file was then imported into Fusion 360 (Autodesk, San Rafael, CA), converted to step format, and all vessels were trimmed.

**Figure 1.**
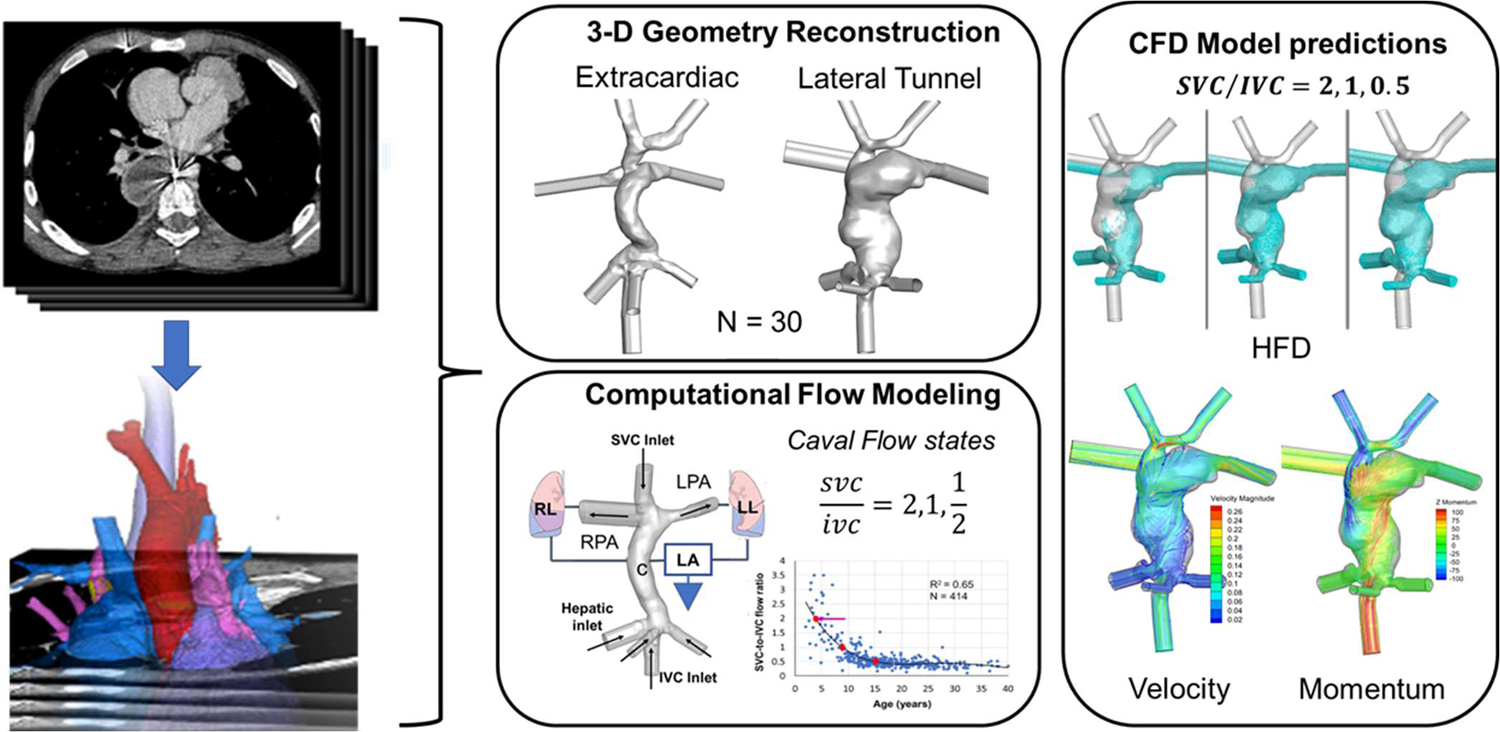
Workflow diagram for CFD-based patient-specific flow analysis

### Computational flow modeling

All patient-specific CFD simulations were carried out using the commercial software ANSYS FLUENT V19.0 and 20.0 (ANSYS, Canonsburg, PA)^26^ with geometries meshed using ANSYS ICEM^26^. CFD model details such as mesh density for adequate flow resolution, solver methodology that ensured solution convergence for each of these models are provided in the Supplementary Material.

The computational flow domain consisted of the patient’s Fontan pathway, SVC (and the bridging vein if present), left and right PAs, IVC, and the main hepatic veins that drained blood into the Fontan-IVC confluence. The hepatic vessels were included in the analysis to capture the realistic local flow dynamic changes at the hepatic-IVC-Fontan anastomosis as hepatic blood interacted with that from IVC at varying flow rates. The inlets (SVC, IVC, hepatic veins) and the outlets (PAs) were extended to ensure numerical stability during flow computation.

Velocities that corresponded to CMR-measured flow rates were directly applied at the SVC inlets. CMR-measured flow rates at the Fontan baffle were split between IVC and hepatic inflow with 25% of total flow from the hepatic vessels based on previous studies^27^. The PA outlets were defined as pressure boundary conditions with a lumped parameter model coupled at the outlets (Figure 1) that compute outlet pressure based on pulmonary vascular resistance (PVR). The pulmonary vascular resistance (PVR) values were individually tuned for each patient CFD model so that the predicted total flow distribution to the LPA and RPA were consistent with CMR measurements within 5% (Supplementary Tables 1 and 2). This ensured consistency between clinically measured total outflow and the model thus allowing for realistic capture of the inflow, outflow and the local flow dynamics within the Fontan pathway to the lungs. Quasi-steady simulations were performed to solve the governing equations (incompressible Navier-Stokes equation) with a Quemada non-Newtonian viscosity model^28^ of blood to capture the effect of hematocrit (45%) and the local shear rate (see Supplementary Materials). HFD and IVC flow distribution, to the lungs were quantified using massless point particles that were uniformly seeded at the hepatic vein and IVC inlets which were passively carried along with the flow to the PA outlets. These particles were tracked, and their flux quantified as they exited the PA outlets ^13, 23^ (see Supplementary Material). For each patient model, CFD simulations based on CMR flow data were performed followed by two additional simulations to represent the age-based changes in SVC and IVC inflows.

### Statistical Analysis

The Shapiro-Wilk test was performed to determine normal distribution of data (See Supplementary data for details). We used the non-parametric Friedman’s chi square test for statistical significance among groups because the predicted data on hepatic flow distribution from our CFD analysis (Figure 3) were not normally distributed and involved three time points (or caval inflow ratios) (See Supplementary Figure 2 for Q-Q plots). Friedman’s test is thought to be more powerful because it makes fewer assumptions than the parametric ANOVA for skewed distributions^29^. A P-value of less than 0.05 indicated a statistically significant difference in hepatic flow distribution between the three age groups. Finally, Pearson correlation coefficient (R) was used to establish the correlation between predicted IVC flow distribution to lungs and HFD for each caval inflow ratio with an assumption that an R > 0.5 represents a strong positive correlation (Detailed description of our statistical methodology is provided in the supplementary material).

## RESULTS

### Impact of age on caval inflows

There were 414 Fontan patients who met inclusion/exclusion criteria and comprised the main retrospective study group. The median age at CMR was 16 [IQR 12, 21] with a range from 2 to 40 years. Supplementary Table 1 details the demographic information of the entire study group. Figure 2A shows the SVC:IVC inflow ratios vs age. SVC:IVC inflow ratios of 2.0, 1.0, and 0.5 corresponded to approximate ages 3, 8, and 15 years old, respectively. There is a non-linear, exponential decline in SVC:IVC inflow ratio from ages 2 to 8, with equal SVC:IVC inflow at around age 8. After age 8, IVC flow predominates but plateaus around age 15 with an SVC:IVC ratio of around 0.5. Accordingly, three simulations were carried out per patient model to predict HFD variation at SVC:IVC ratios of 2.0, 1.0, and 0.5, also adjusting for an estimated age-based corresponding change in total flow^30–32^. Given that most Fontan procedures occur between ages 3-5 years, this change in SVC:IVC inflow ratio may explain some of the changes in HFD over time (see below). Figure 2B describes indexed total caval inflow (SVC + IVC) indexed to BSA vs age which tends to have less changes as patients get older.

**Figure 2.**
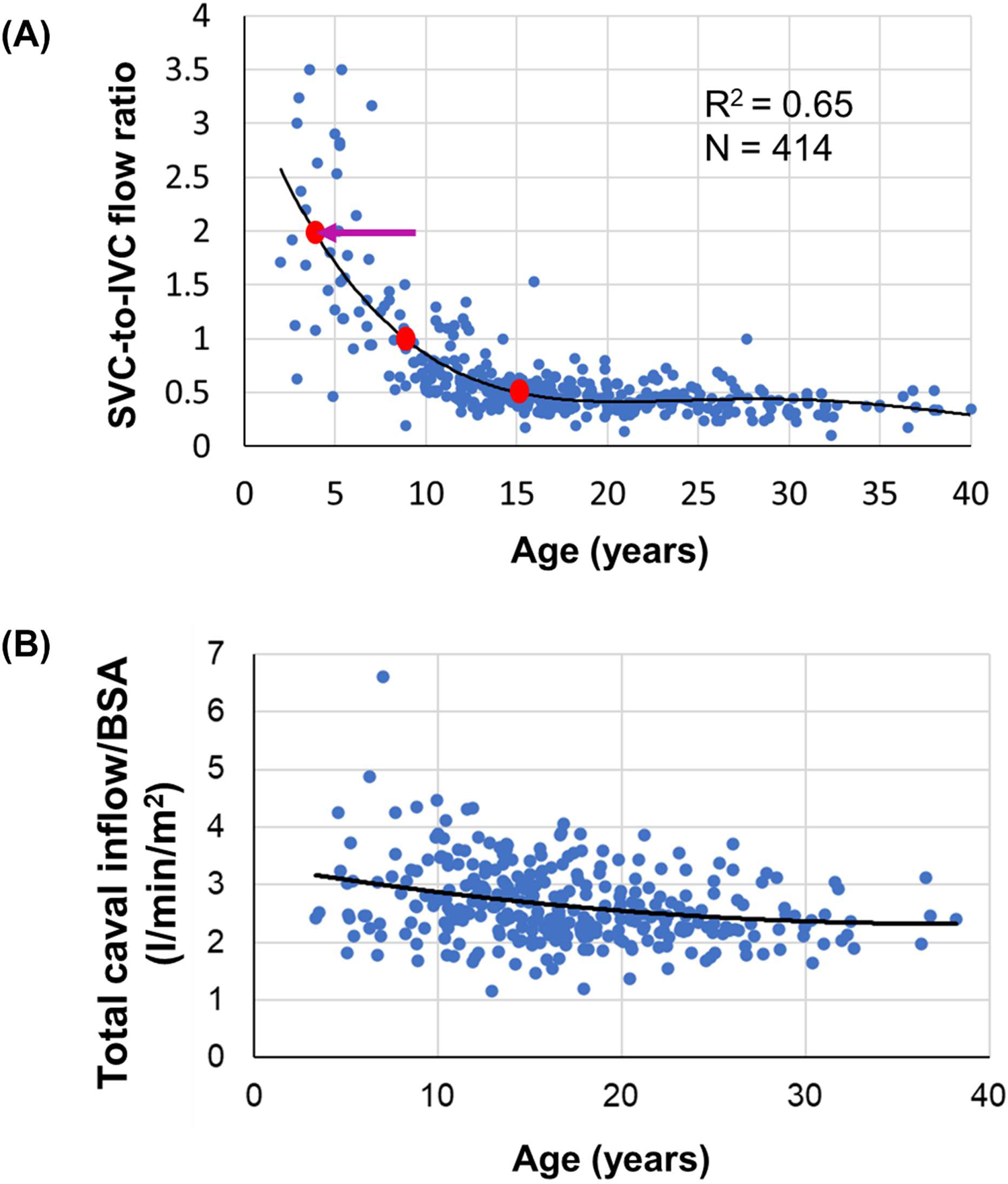
SVC-IVC Analysis based on CMR data from 414 Fontan patients (A) shows the shift in caval inflow ratio with age. Red dots indicate the flow ratios in which patient-specific CFD simulations were performed. Arrow indicates the typical time of Fontan completion in single ventricle patients; (B) Shows total caval inflow indexed to BSA vs age.

### Impact of change in caval inflow on hepatic flow distribution

CFD predictions showed that variations of HFD in response to changes in caval inflow were different in both EC and LT cohorts. In patients with an EC Fontan (n=30), the median CFD-predicted differential pulmonary artery blood flow percentage to the right/left lungs was 54/46%, 53/47%, and 53/47% for caval SVC:IVC ratios of 2.0, 1.0, and 0.5 respectively (Figure 3A). Median differential HFD percentage to right/left lungs were 70/30%, 66/34%, and 65/35% for caval ratios of 2.0, 1.0, and 0.5, respectively (Figure 3B). There was no significant change in pulmonary artery blood flow distribution or HFD (P=0.46) with changes in age or SVC:IVC flow in patients with an EC Fontan conduit.

**Figure 3.**
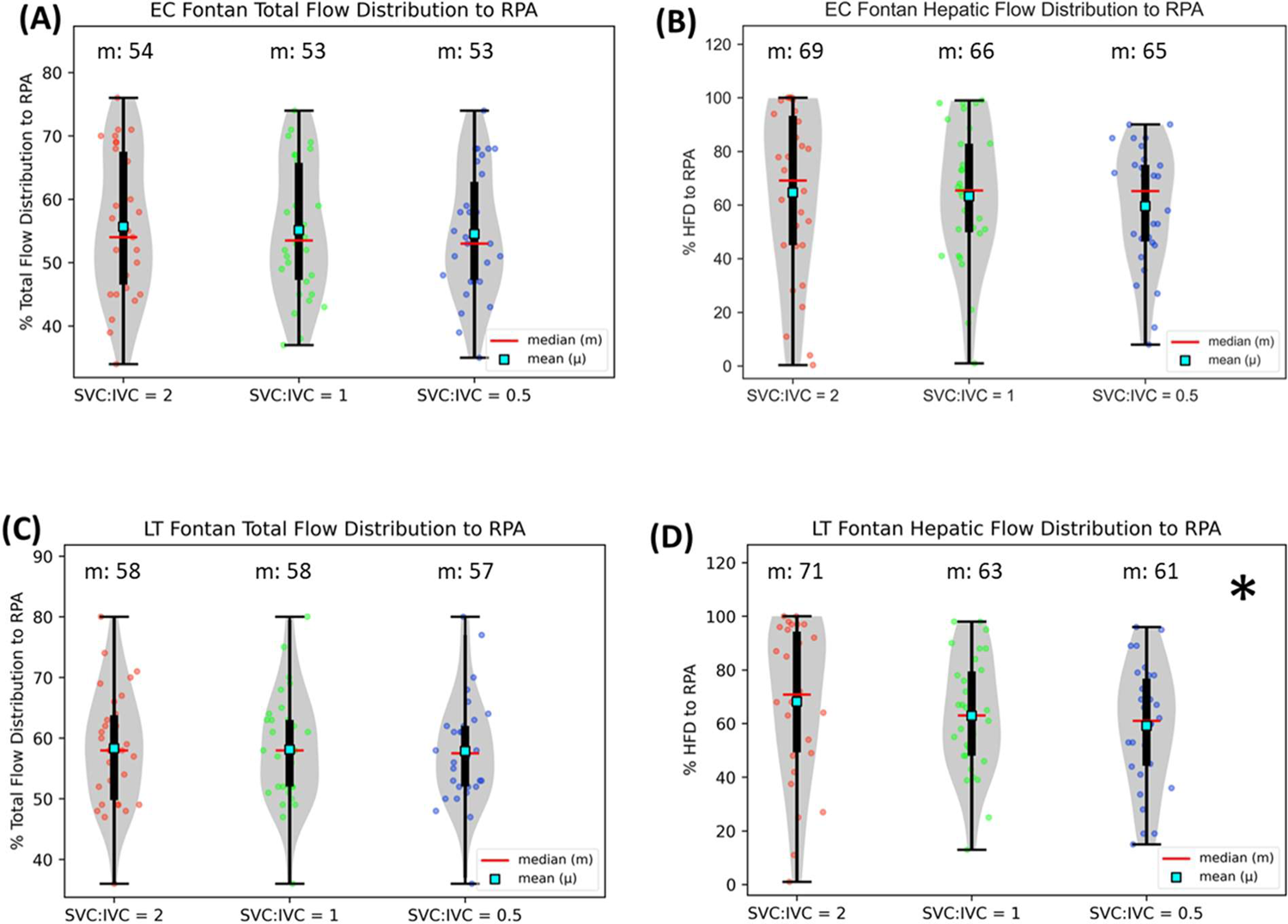
Variation in total flow distribution and hepatic flow distribution in our (A & B) extracardiac Fontan cohort (n=30) and (C & D) lateral tunnel cohort (n=30) as caval flow ratio shifts with age. Note that figure shows HFD to RPA (LPA = 1-RPA).* indicates statistical significance

For patients with a LT Fontan (n=30), CFD-predicted median differential pulmonary artery blood flow percentage to the right/left lungs was 58/42%, 58/42%, and 57/43% for caval ratios of 2.0, 1.0, and 0.5 respectively (Figure 3C). Median differential HFD percentage to right/ left lungs were 71/29%, 63/27%, and 61/39% for SVC:IVC caval ratios of 2.0, 1.0, and 0.5, respectively (Figure 3D). The change in HFD distribution across three caval inflow ratios was statistically significant (P=0.04). There were no other significant changes in pulmonary artery blood flow in patients with a LT Fontan conduit. These data suggest that the increased proportion of IVC (vs SVC) inflow are driving changes in HFD in patients with a LT Fontan, but not patients with an EC Fontan.

### Strength of positive correlation between IVC flow distribution to PAs and HFD moderately decreases with age

Twenty-six patients (13 EC, 13 LT) randomly selected from our EC and LT cohorts were further analyzed to determine the correlation between IVC flow distribution to branch PAs and HFD to branch PAs. Strong positive correlations existed between HFD vs IVC flow distribution of r=0.86 (P<0.0001), r=0.73 (P<0.0001), and r=0.63 (P<0.001) for SVC:IVC ratios of 2.0, 1.0, and 0.5, respectively (Figure 4). The strength of the correlation decreased slightly over increased IVC flow (or age). Specific patient examples of momentum analysis with graphical depictions are shown and described in Figures 5-8. The greater momentum of the SVC flow at age ∼3 (left panels of Figures 5-8 A), seemed to preferentially divert the IVC flow that seemed to carry the hepatic blood flow (left panels of Figures 5-8 B and C). This flow phenomenon may be attributed to the strong positive correlation between HFD and IVC for SVC:IVC ratio of 2. As IVC flow increases with age (middle and right panels of Figures 5-8 A), flow disturbance induced by the increased momentum from IVC flow at the IVC-hepatic anastomosis and eventually competing with SVC at Fontan-PA-SVC anastomosis may have contributed to the slight decrease in correlation between HFD and IVC for SVC:IVC ratios of 2 and 0.5 (middle and right panels of Figures 5-8 B and C, Figure 4 B and C).

**Figure 4.**
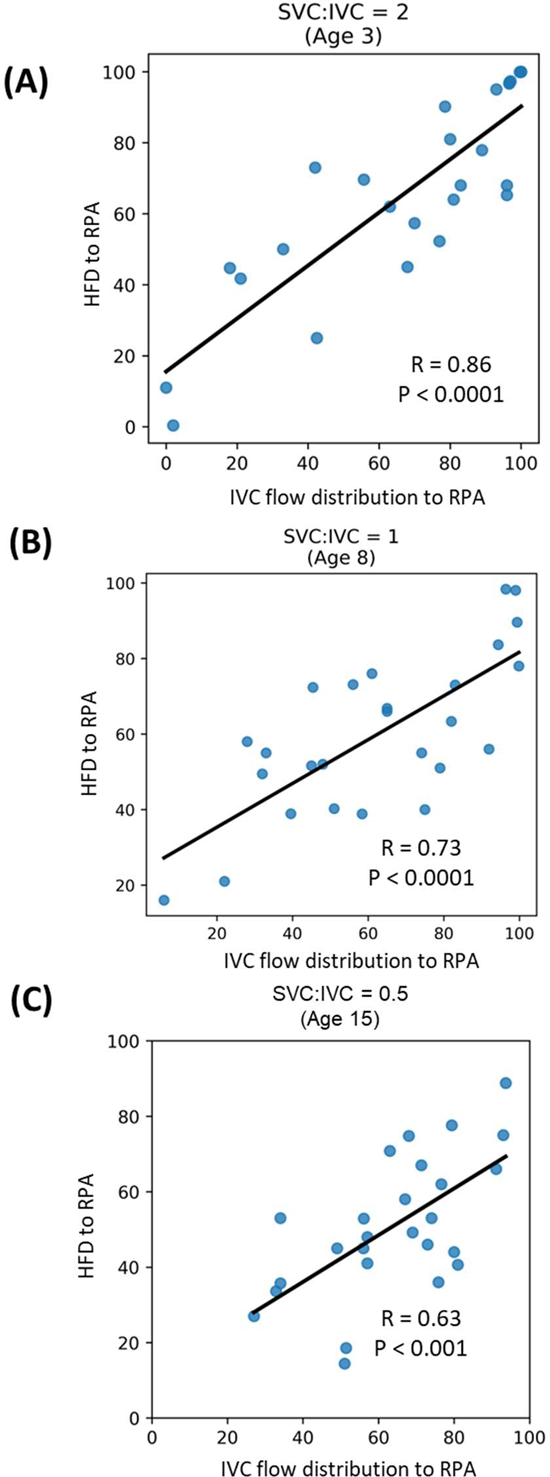
Correlation analysis between IVC flow distribution and HFD to RPA was performed using 26 patient CFD models (13 EC + 13 LT) to determine the Pearson correlation coefficient (ρ) for each of the SVC:IVC flow ratios (A-C)

**Figure 5.**
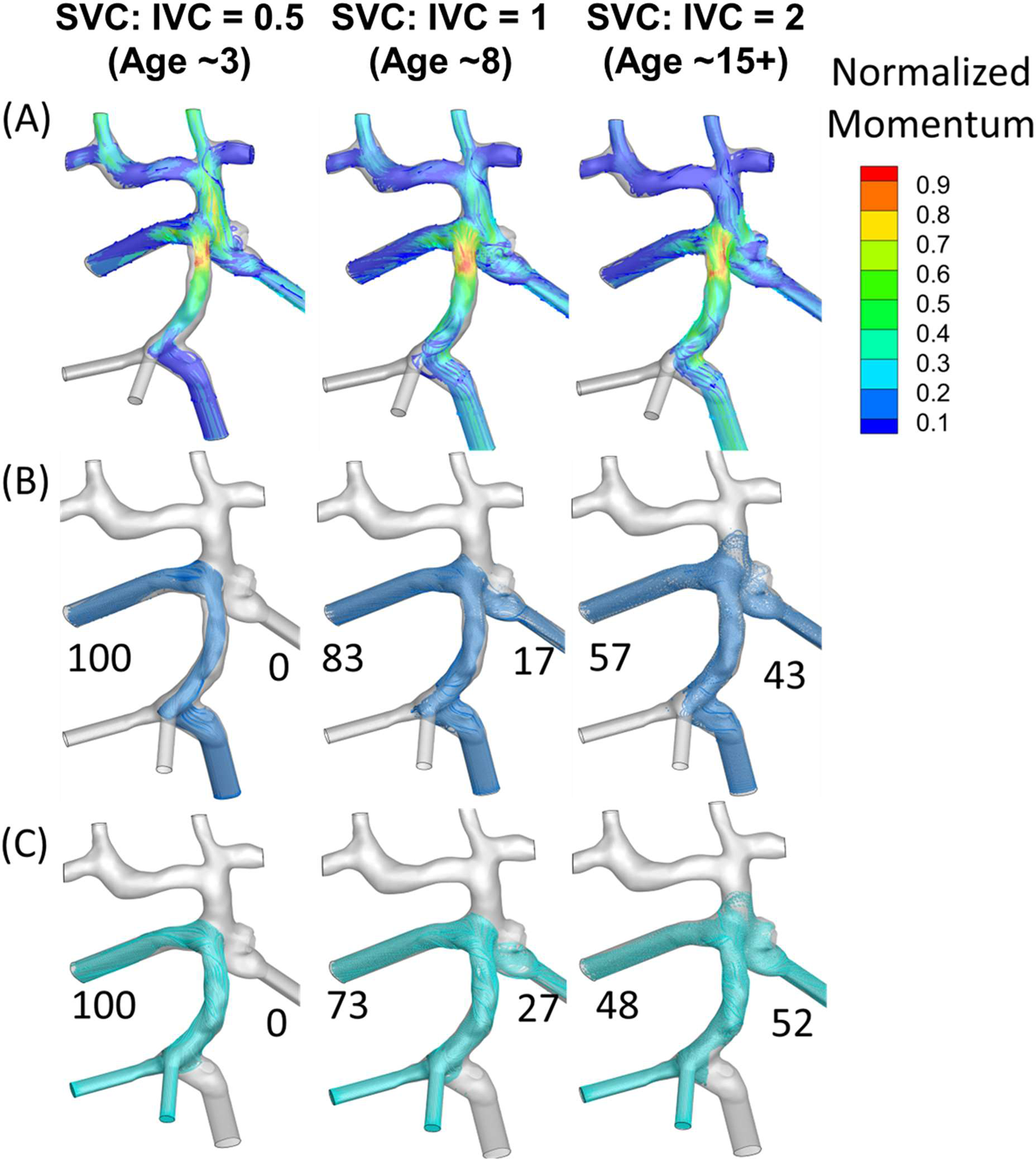
Momentum analysis on Patient 13 Extracardiac Fontan pathway (A): Contour plots of normalized (to maximum) momentum; (B) % IVC flow distribution to lungs; and (C) % Hepatic flow distribution to lungs

**Figure 6.**
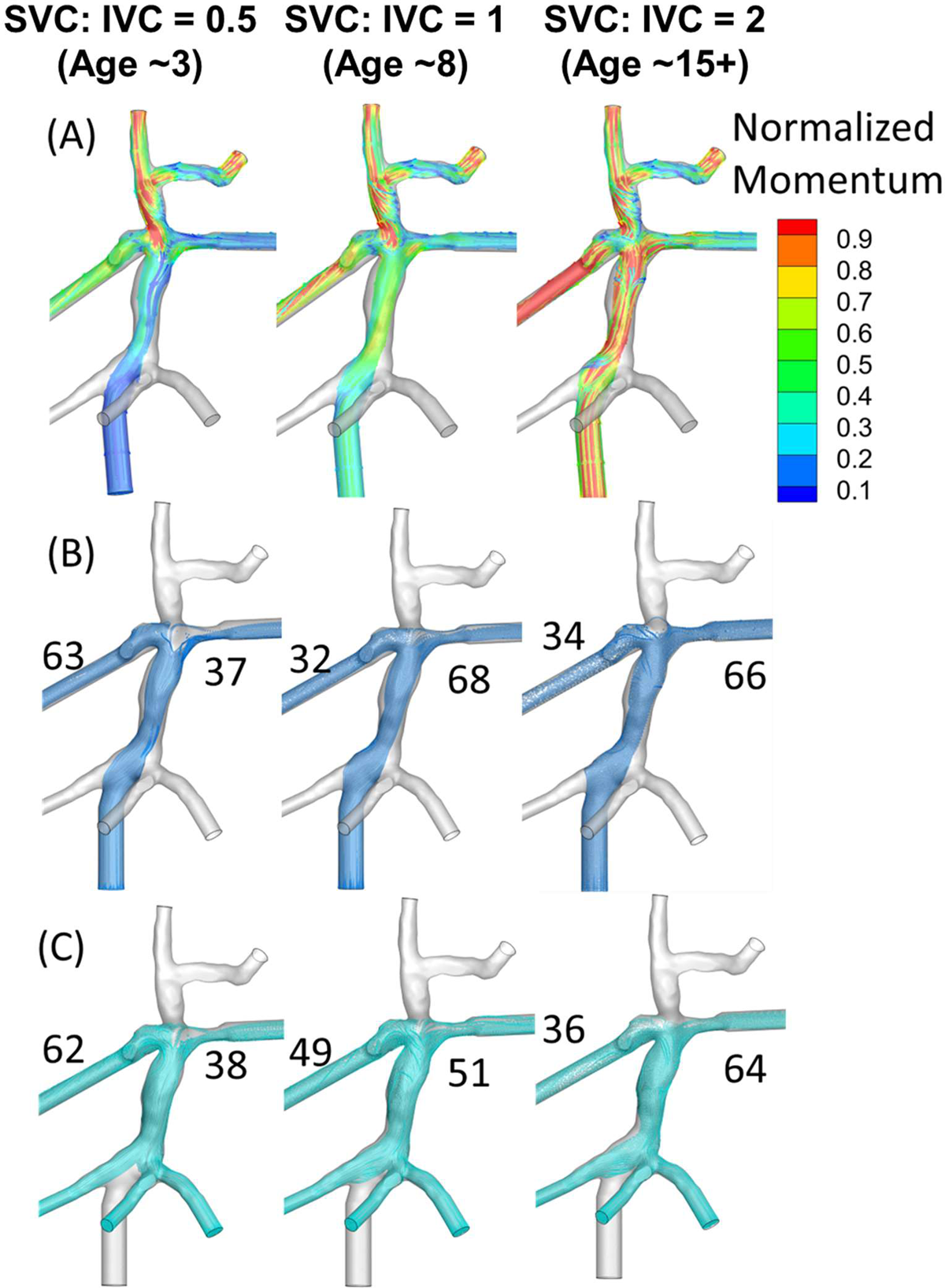
Momentum analysis on Patient 16 Extracardiac Fontan pathway (A): Contour plots of normalized (to maximum) momentum; (B) % IVC flow distribution to lungs; and (C) % Hepatic flow distribution to lungs

**Figure 7.**
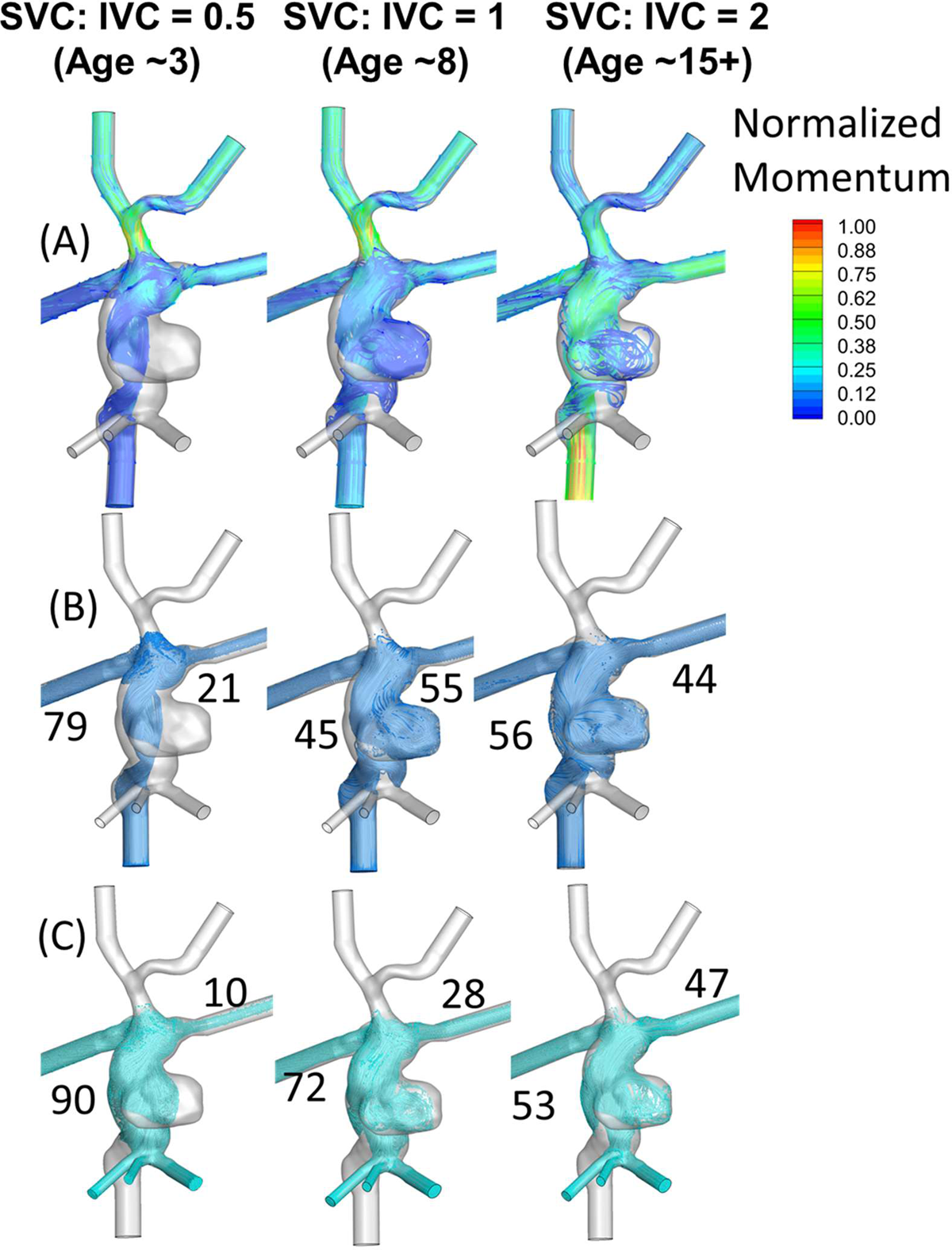
Momentum analysis on Patient 13 Lateral tunnel Fontan pathway (A): Contour plots of normalized (to maximum) momentum; (B) % IVC flow distribution to lungs; and (C) % Hepatic flow distribution to lungs

**Figure 8.**
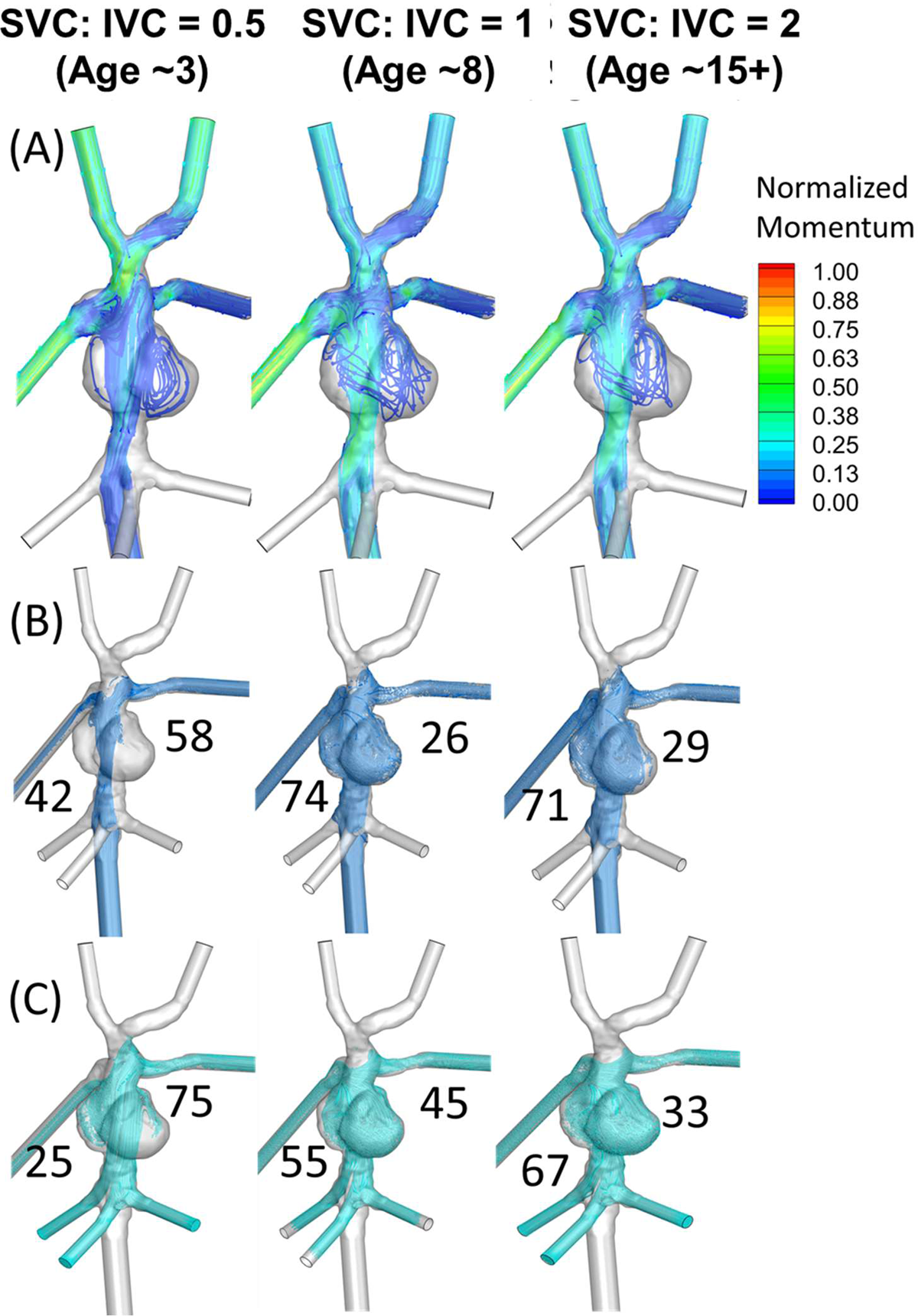
Momentum analysis on Patient 30 Lateral tunnel Fontan pathway (A): Contour plots of normalized (to maximum) momentum; (B) % IVC flow distribution to lungs; and (C) % Hepatic flow distribution to lungs

## Discussion

Blood flow and its interaction with its lumen is a dynamic process that changes with any perturbation caused by either age, growth, disease, or intervention. These hemodynamic concepts are also relevant for the Fontan circulation. With the improved survival of Fontan patients leading and an increase in the worldwide population of surviving Fontan patients with various types of single ventricle anatomy^5^, a broader understanding of how Fontan hemodynamics alters with age can help with the management and prevention of future complications^12, 14–16, 22, 23^. This study establishes the quantitative shift from SVC dominant to IVC dominant flow as Fontan patients age with an inversion point around age 8. In younger patients with a higher SVC:IVC ratio (2.0), both EC and LT Fontan patients had mild maldistribution of HFD (Figure 3A and C). Patients with a LT had a more favorable “correction” with increased IVC flow (right HFD decreased from 71% ◊ 61%) (Figure 3D). Patients with a EC Fontan did not seen this correction and remained steady across the various flow condition and with increasing age (Figure 3B). This study is the first to show that long-term variations in HFD are impacted by the type of Fontan procedure.

Results from this study show how SVC:IVC inflows into the Fontan pathway change with age and how those changes impact the local hemodynamics and HFD. CMR data collected from a large (n=414) group of Fontan patients revealed a curvilinear change in SVC and IVC flow ratio with age (2/3^rd^ SVC at age∼3 transitioning to 1/3^rd^ SVC at age 15+), indicating that the Fontan inflows can be dynamic at least until adulthood upon which IVC flow dominates (Figure 2). These data are highly consistent with a previous Doppler study by Salim et al., that showed that SVC flow decreased (to 35%) as patients got older^33^. Our CFD model predictions showed that patients with a LT Fontan were more likely to have maldistribution of HFD when SVC inflow dominates (age ∼3). This is clinically significant as imbalances to HFD can lead to PAVMs. This maldistribution improved with CFD models with increased IVC flow. EC Fontan physiology was generally more favorable with more symmetric HFD early in life and more consistent even with changes in SVC:IVC flow ratios. These data reinforce the importance of growth-related changes in physiology that should be incorporated while predicting any long-term Fontan hemodynamics.

While it is desirable to have a balanced HFD to prevent PAVMs in Fontan patients, a minimum level of hepatic factor required to prevent PAVMs has not been quantitatively established. Many studies have shown that a complete lack of hepatic blood effluence led to the formation of PAVM^34–36^. At the same time, there is clinical evidence showing complete resolution of PAVM following redirection of hepatic venous blood to the affected lung^6, 7, 11, 37^. In our patient cohort, while many patients had a balanced HFD (Figure 3), there were some patients with highly skewed HFD especially when SVC inflow dominated the Fontan inflows (age ∼3). Our model predictions suggested that HFD tends to achieve some balance with age for these patients consistent with other patient models. These data highlight patient-specificity, how SVC-IVC caval inflow change with respect to age can impact HFD, and the need for personalized planning of Fontan pathways.

Long term outcomes of extracardiac vs lateral tunnel Fontan procedures has been a subject of continuous debate^5, 15^ with most decision making highly influenced by individual surgeon choice and institutional preferences^5^. A comprehensive evaluation of power loss utilizing by Haggerty et al^15^, revealed no considerable differences between the connection types. The current study provides new quantitative insights with respect to HFD and introduces some concern for early maldistribution of HFD in both Fontan types, but improved balance in patients with a LT Fontan.

Dasi et al identified the caval-offset induced momentum barrier created by the SVC flow being the main factor for unbalanced HFD in extracardiac Fontan^12^. These findings are highly consistent with our results, especially for age ∼3 when SVC flow dominated (Figures. 5-8). Also, the highest skewness in HFD occurred for an SVC-IVC flow ratio of 2 (Age ∼3) (Figure. 3) for both EC and LT cohorts highlighting the impact of SVC. Furthermore, our momentum analysis revealed that the potential long-term impact of the SVC momentum barrier, while significant immediately after Fontan completion (Age ∼3) diminishes as IVC flow dominated with patient growth (age 8 and 15+, Figures 5-8). These results demonstrate how competing momentum between in the Fontan pathway impacts HFD during the course of time.

Our study has the following limitations. All the simulations were performed using time-averaged flow conditions measured from the MRI with rigid wall assumptions. Rigid wall assumptions were made based on the conclusions from previous study that there was no significant change in HFD due to wall deformation^38^. The growth of PAs was not accounted for based on the study that reported nonsignificant growth of vessels commensurate with increasing flow^21, 39^. Fontan flow is known to be transient (e.g.: impact of respiration) and hence, the time-average flow conditions may introduce small quantitative differences. The potential growth and geometry size changes of the LT baffle that can occur with time were also not able to be estimated in this study. Finally, a constant PVR was assumed for all the SVC-to-IVC caval flow ratios while PVR can potentially change over time.

In conclusion, this study described the SVC:IVC flow ratios in a large cohort of patients ranging from 2 to 40 years old, showing the conversion of SVC dominant flow to IVC dominant flow when patients are approximately 8 years old. Using CMR-based patient-specific CFD modeling, both EC and LT Fontan patients had a tendency towards maldistribution of HFD early in line. With increasing IVC inflow with age, patients with a LT had improved HFD distribution, while EC Fontan patients had a relatively consistent HFD. Our analysis provided quantitative insights into the impact of changing caval inflows on long-term hepatic flow distribution in extracardiac and lateral tunnel Fontan types. These data also showed the importance of incorporating SVC:IVC changes over time in CFD modeling toward gaining insights into for long-term hemodynamics of Fontan.

## Supplementary Data

Supplementary Methods contains detailed description of methodology used in computational flow modeling and statistical analysis.

Supplementary table 1: Demographic details of the extracardiac patient cohort considered in our study. Also shown are IVC, SVC, MRI measured- and CFD predicted flow split.

Supplementary table 2: Demographic details of the lateral tunnel patient cohort considered in our study. Also shown are IVC, SVC, MRI measured- and CFD predicted flow split.

## Disclosure

### Author Contributions

VG conceptualized the study and wrote the manuscript. VG and RR designed the study. LM, EE, JD, MC, and NS performed the segmentation and reconstruction of the CMR data. VG, AS, and MC developed the computational models and curated the data. N St.C performed data collection and curation. All authors analyzed the results, reviewed, and edited the manuscript.

## Data Availability

80 CMR-based patient-specific computational flow modeling simulations were performed to predict the hepatic flow distribution HFD) using 30 extracardiac and 30 Lateral tunnel Fontan [(30+30)*3 flow conditions = 180). The HFD data was used in our analysis to derive the quantitative insights and statistical analysis presented in our study. We have also generated the HFD plots for each of the patient specific models (total = 180) that can provide spatial insights into how hepatic flow gets distributed for a given patient Fontan pathway and caval inflow. While a few examples are provided in our manuscript, we will be more than happy to provide all the plots as supplementary figures upon recommendation from the editors and reviewers.

## Acknowledgements

Research reported in this publication was supported by the National Heart, Lung, And Blood Institute of the National Institutes of Health under Award Number R01HL161507. The content is solely the responsibility of the authors and does not necessarily represent the official views of the National Institutes of Health. Authors also acknowledge the computational resources provided by the Texas Advanced Computer Center for performing the flow simulations as a part of this study.

## References

1. Mazza GA, Gribaudo E and Agnoletti G. The pathophysiology and complications of Fontan circulation. Acta Bio Medica: Atenei Parmensis. 2021;92.

2. Kavarana MN, Jones JA, Stroud RE, Bradley SM, Ikonomidis JS and Mukherjee R. Pulmonary arteriovenous malformations after the superior cavopulmonary shunt: mechanisms and clinical implications. Expert review of cardiovascular therapy. 2014;12:703–713.

3. Maxey TS, Herlong JR, Jansen LN and Kirshbom PM. Fontan Procedure *Critical Heart Disease in Infants and Children*: Elsevier; 2019: 747–757. e2.

4. Cartin-Ceba R, Swanson KL and Krowka MJ. Pulmonary arteriovenous malformations. Chest. 2013;144:1033–1044.

5. Rychik J, Atz AM, Celermajer DS, Deal BJ, Gatzoulis MA, Gewillig MH, Hsia T-Y, Hsu DT, Kovacs AH and McCrindle BW. Evaluation and management of the child and adult with Fontan circulation: a scientific statement from the American Heart Association. Circulation. 2019;140:e234–e284.

6. Pike NA, Vricella LA, Feinstein JA, Black MD and Reitz BA. Regression of severe pulmonary arteriovenous malformations after Fontan revision and “hepatic factor” rerouting. The Annals of thoracic surgery. 2004;78:697–699.

7. Wu I-H and Nguyen KH. Redirection of hepatic drainage for treatment of pulmonary arteriovenous malformations following the Fontan procedure. Pediatric cardiology. 2006;27:519–522.

8. Srivastava D, Preminger T, Lock JE, Mandell V, Keane JF, Mayer Jr JE, Kozakewich H and Spevak PJ. Hepatic venous blood and the development of pulmonary arteriovenous malformations in congenital heart disease. Circulation. 1995;92:1217–1222.

9. Spearman AD and Ginde S. Pulmonary Vascular Sequelae of Palliated Single Ventricle Circulation: Arteriovenous Malformations and Aortopulmonary Collaterals. Journal of Cardiovascular Development and Disease. 2022;9:309.

10. Yang W, Vignon-Clementel IE, Troianowski G, Reddy VM, Feinstein JA and Marsden AL. Hepatic blood flow distribution and performance in conventional and novel Y-graft Fontan geometries: a case series computational fluid dynamics study. The Journal of thoracic and cardiovascular surgery. 2012;143:1086–1097.

11. Sundareswaran KS, de Zélicourt D, Sharma S, Kanter KR, Spray TL, Rossignac J, Sotiropoulos F, Fogel MA and Yoganathan AP. Correction of pulmonary arteriovenous malformation using image-based surgical planning. JACC: Cardiovascular Imaging. 2009;2:1024–1030.

12. Dasi LP, Whitehead K, Pekkan K, de Zelicourt D, Sundareswaran K, Kanter K, Fogel MA and Yoganathan AP. Pulmonary hepatic flow distribution in total cavopulmonary connections: extracardiac versus intracardiac. The Journal of thoracic and cardiovascular surgery. 2011;141:207–214.

13. De Zélicourt DA, Haggerty CM, Sundareswaran KS, Whited BS, Rossignac JR, Kanter KR, Gaynor JW, Spray TL, Sotiropoulos F and Fogel MA. Individualized computer-based surgical planning to address pulmonary arteriovenous malformations in patients with a single ventricle with an interrupted inferior vena cava and azygous continuation. The Journal of thoracic and cardiovascular surgery. 2011;141:1170–1177.

14. Whitehead KK, Pekkan K, Kitajima HD, Paridon SM, Yoganathan AP and Fogel MA. Nonlinear power loss during exercise in single-ventricle patients after the Fontan: insights from computational fluid dynamics. Circulation. 2007;116:I-165–I-171.

15. Haggerty CM, Restrepo M, Tang E, de Zélicourt DA, Sundareswaran KS, Mirabella L, Bethel J, Whitehead KK, Fogel MA and Yoganathan AP. Fontan hemodynamics from 100 patient-specific cardiac magnetic resonance studies: a computational fluid dynamics analysis. The Journal of thoracic and cardiovascular surgery. 2014;148:1481–1489.

16. Khiabani RH, Whitehead KK, Han D, Restrepo M, Tang E, Bethel J, Paridon SM, Fogel MA and Yoganathan AP. Exercise capacity in single-ventricle patients after Fontan correlates with haemodynamic energy loss in TCPC. Heart. 2015;101:139–143.

17. Wang Q, Primiano C, McKay R, Kodali S and Sun W. CT image-based engineering analysis of transcatheter aortic valve replacement. JACC: Cardiovascular Imaging. 2014;7:526–528.

18. Bossers SS, Cibis M, Gijsen FJ, Schokking M, Strengers JL, Verhaart RF, Moelker A, Wentzel JJ and Helbing WA. Computational fluid dynamics in Fontan patients to evaluate power loss during simulated exercise. Heart. 2014;100:696–701.

19. Cibis M, Jarvis K, Markl M, Rose M, Rigsby C, Barker AJ and Wentzel JJ. The effect of resolution on viscous dissipation measured with 4D flow MRI in patients with Fontan circulation: Evaluation using computational fluid dynamics. Journal of biomechanics. 2015;48:2984–2989.

20. Bove E, De Leval M, Migliavacca F, Balossino R and Dubini G. Toward optimal hemodynamics: computer modeling of the Fontan circuit. Pediatric cardiology. 2007;28:477–481.

21. Restrepo M, Tang E, Haggerty CM, Khiabani RH, Mirabella L, Bethel J, Valente AM, Whitehead KK, McElhinney DB and Fogel MA. Energetic implications of vessel growth and flow changes over time in Fontan patients. The annals of thoracic surgery. 2015;99:163–170.

22. Wei Z, Singh-Gryzbon S, Trusty PM, Huddleston C, Zhang Y, Fogel MA, Veneziani A and Yoganathan AP. Non-Newtonian effects on patient-specific modeling of Fontan hemodynamics. Annals of biomedical engineering. 2020;48:2204–2217.

23. Trusty PM, Wei ZA, Slesnick TC, Kanter KR, Spray TL, Fogel MA and Yoganathan AP. The first cohort of prospective Fontan surgical planning patients with follow-up data: How accurate is surgical planning? The Journal of thoracic and cardiovascular surgery. 2019;157:1146–1155.

24. van Bakel TM, Lau KD, Hirsch-Romano J, Trimarchi S, Dorfman AL and Figueroa CA. Patient-specific modeling of hemodynamics: supporting surgical planning in a Fontan circulation correction. Journal of cardiovascular translational research. 2018;11:145–155.

25. Frieberg P, Aristokleous N, Sjöberg P, Töger J, Liuba P and Carlsson M. Computational fluid dynamics support for fontan planning in minutes, not hours: the next step in clinical pre-interventional simulations. Journal of cardiovascular translational research. 2022;15:708–720.

26. Guide AFT. ANSYS fluent tutorial Guide 18. ANSYS Fluent Tutorial Guide 18. 2018;15317:724–746.

27. Granger DN and Kvietys PR. Circulation, overview. 2004.

28. Quemada D. Rheology of concentrated disperse systems II. A model for non-newtonian shear viscosity in steady flows. Rheologica Acta. 1978;17:632–642.

29. Pereira DG, Afonso A and Medeiros FM. Overview of Friedman’s test and post-hoc analysis. Communications in Statistics-Simulation and Computation. 2015;44:2636–2653.

30. Cattermole GN, Leung PM, Mak PS, Chan SS, Graham CA and Rainer TH. The normal ranges of cardiovascular parameters in children measured using the Ultrasonic Cardiac Output Monitor. Critical care medicine. 2010;38:1875–1881.

31. Yoganathan AP, Fogel M, Gamble S, Morton M, Schmidt P, Secunda J, Vidmar S and Del Nido P. A new paradigm for obtaining marketing approval for pediatric-sized prosthetic heart valves. The Journal of thoracic and cardiovascular surgery. 2013;146:879–886.

32. Mannarino S, Bulzomì P, Codazzi AC, Rispoli GA, Tinelli C, De Silvestri A, Manzoni F and Chiapedi S. Inferior vena cava, abdominal aorta, and IVC-to-aorta ratio in healthy Caucasian children: Ultrasound Z-scores according to BSA and age. Journal of Cardiology. 2019;74:388–393.

33. Salim MA, DiSessa TG, Arheart KL and Alpert BS. Contribution of superior vena caval flow to total cardiac output in children: a Doppler echocardiographic study. Circulation. 1995;92:1860–1865.

34. Shah MJ, Rychik J, Fogel MA, Murphy JD and Jacobs ML. Pulmonary AV malformations after superior cavopulmonary connection: resolution after inclusion of hepatic veins in the pulmonary circulation. The Annals of thoracic surgery. 1997;63:960–963.

35. Ashrafian H and Swan L. The mechanism of formation of pulmonary arteriovenous malformations associated with the classic Glenn shunt (superior cavopulmonary anastomosis). Heart. 2002;88:639–639.

36. Vettukattil J. Pathogenesis of pulmonary arteriovenous malformations: role of hepatopulmonary interactions. 2002;88:561–563.

37. McElhinney DB, Marx GR, Marshall AC, Mayer JE and Del Nido PJ. Cavopulmonary pathway modification in patients with heterotaxy and newly diagnosed or persistent pulmonary arteriovenous malformations after a modified Fontan operation. The Journal of Thoracic and Cardiovascular Surgery. 2011;141:1362–1370. e1.

38. Long C, Hsu MC, Bazilevs Y, Feinstein J and Marsden A. Fluid–structure interaction simulations of the Fontan procedure using variable wall properties. International journal for numerical methods in biomedical engineering. 2012;28:513–527.

39. Restrepo M, Mirabella L, Tang E, Haggerty CM, Khiabani RH, Fynn-Thompson F, Valente AM, McElhinney DB, Fogel MA and Yoganathan AP. Fontan pathway growth: a quantitative evaluation of lateral tunnel and extracardiac cavopulmonary connections using serial cardiac magnetic resonance. The Annals of thoracic surgery. 2014;97:916–922.

